# Kidney Damage and Associated Risk Factors in the Rural Eastern Cape, South Africa: A Cross-Sectional Study

**DOI:** 10.1101/2023.09.21.23295885

**Authors:** Ernesto Rosales Gonzalez, Parimalanie Yogeswaran, Jimmy Chandia, Guillermo Alfredo Pulido Estrada, Oladele Vincent Adeniyi

## Abstract

**Background:** The colliding epidemic of infectious and non-communicable diseases in South Africa could potentially increase the prevalence of kidney disease. This study determines the prevalence of kidney damage and known risk factors in a rural community of the Eastern Cape province, South Africa.

**Methods:** This observational cross-sectional study was conducted in the outpatient department of the Mbekweni Community Health Centre in the Eastern Cape between May and July 2022. Relevant data on demography, medical history, anthropometry and blood pressure were obtained. The glomerular filtration rate was estimated using the Chronic Kidney Disease Epidemiology Collaboration Creatinine (CKD-EPI_Creatinine_) equation and the re-expressed four-variable Modification of Diet in Renal Disease (MDRD) equation, without any adjustment for black ethnicity. Significant kidney damage was defined as low eGFR (<60mL/min per 1.73m^2^) and/or the presence of proteinuria. We used the logistic regression model analysis to identify the independent risk factors for significant kidney damage.

**Results:** The mean (±standard deviation) age of the 389 participants was 52.3 (± 17.5) years. The prevalence of significant kidney damage was 17.2% (n=67), as estimated by the CKD-EPI_Creatinine,_ with slight difference from the MDRD equation (17.7%; n=69), while the prevalence of proteinuria was 7.2%. Risk factors for significant kidney damage were older age (OR=0.94, 95% CI 0.91 - 0.96, p<0.001) and the presence of proteinuria (OR=0.98, 95% CI 0.97 - 0.99, p 0.002). Proteinuria was strongly associated with hypertension (OR=4.46, 95% CI 1.33 - 14.92, p<0.015) and elevated serum creatinine (OR=1.01, 95% CI 1 - 1.02, p=0.004).

Conclusions

This study found a high prevalence of kidney damage (17.2%) and proteinuria (7.2%) in this rural community, largely attributed to advanced age and hypertension, respectively. Early detection of proteinuria and decreased renal function could lead to prompt preventative measures and management to delay the progression to end-stage kidney failure and mortality.

## Introduction

Chronic kidney disease (CKD) is a growing problem in developing countries [1,2]. In most of sub-Saharan Africa, most patients with CKD die because of a lack of adequate treatment and renal replacement therapy (RRT) [3]. Renal replacement therapy (RRT) is very expensive, making it unaffordable to people in low- and middle-income countries. Evidence suggests an increase in the demand for renal replacement therapy in South Africa, from 70 per million of the population in 1994 to 190 per million of the population in 2017, which is a more than two-fold increase [4,5]. The true burden of chronic kidney disease in South Africa is unknown, owing to a lack of nationally representative studies in the country. Evidence suggests an increasing burden of non-communicable diseases in South Africa (hypertension, obesity and Type-2 diabetes mellitus) which are known risk factors for end-stage kidney disease (ESKD), particularly in black ethnic groups [6–8].

CKD often goes unnoticed, because there are no specific symptoms, leading to delays in diagnosis or diagnosis at an advanced stage. Investigations for CKD are very simple and freely available in South Africa. The diagnosis of CKD is made in the presence of a persistently low glomerular filtration rate (GFR), which is estimated from the serum creatinine concentration or proteinuria over a period of at least three months. Evidence shows unequivocally that the Chronic Kidney Disease Epidemiology Collaboration Creatinine (CKD-EPI_Creatinine_) equation is more accurate than the previous Modification of Diet for Renal Disease equation [2,9]. The diagnostic challenges of CKD cannot be ignored, especially in rural communities where timeous access to relevant tests remains a concern. It has been estimated that 10% of the world’s population has some degree of CKD, with trends suggesting an alarming increase in the incidence globally [10]. Current data estimates that about 5 million South Africans over the age of 20 years have CKD, and in black South Africans, the figure is almost certainly higher [3,10]. In sub-Saharan Africa, the overall prevalence in the general population is 15,8 % for CKD Stages 1–3 and 4.6% for Stages 3–5 in the general population [11].

CKD complications represent a considerable burden on global healthcare resources, and only a few countries have sufficiently robust economies to meet the challenge posed by this disease [1]. As such, an intensified screening programme aimed at prioritising at-risk individuals at the population level might assist in early diagnosis and management to prevent further progression to end-stage kidney failure and mortality [4]. Without a nationally representative study sample, it is difficult to gain a full understanding of the true burden of CKD in the country. Worse still, there is a paucity of published literature on CKD among people living in rural communities, especially in the Eastern Cape province. This has serious implications for crafting an effective national health promotion and disease prevention policy for the country. In order to contribute much-needed data on the burden of CKD among rural residents, which is currently unavailable in the country, this study determined the prevalence of kidney damage and examined the associated risk factors among individuals accessing care at the rural Mbekweni Community Health Centre in the Eastern Cape province, South Africa.

## Methods

### Ethical considerations

This study received ethical approval from the Research Ethics Committee of the Faculty of Medicine and Health Sciences, Walter Sisulu University (Reference Number: 025/2017). Permission for the study’s implementation was granted by the Eastern Cape Department of Health, OR Tambo District Department of Health and the facility manager. Each participant gave written informed consent, indicating voluntary participation in the study. The study was implemented in accordance with the Helsinki Declaration and Good Clinical Practice Guidelines.

### Study design, setting and population

This observational cross-sectional study was conducted in the outpatient department of the Mbekweni Community Health Centre (CHC) in the King Sabata Dalindyebo (KSD) sub-district of OR Tambo district in the Eastern Cape between May and July 2022. This CHC serves the deeply rural residents of Mbekweni location, comprising an estimated population of 24 284 inhabitants [12]. The adult population (20 years and above) of Mbekweni is estimated to be 13 716 [12]. The sample size of 389 was estimated by using the free Software Epidat 3.1. Xunta de Galicia, Spain (Pan-American Health Organization/World Health Organization), using a confidence level of 95%, an expected proportion of 10%, and a maximum error of 3%.

In order to ensure inclusivity and minimise selection bias, participants were recruited through a simple random sampling technique; a hundred numbers were given to the adults attending the outpatient department of Mbekweni CHC, and 50 numbers were chosen randomly using the free Software Epidat 3.1. Xunta de Galicia Spain (Pan-American Health Organization/World Health Organization) run on Dr ERG’s personal computer.

Participants were considered eligible if they were ≥ 20 years at the time of the study, attending the outpatient department of Mbekweni CHC, and willing to participate in the research study. However, participants were excluded if they had acute medical (vomiting, diarrhoea, burns) or psychiatric emergencies that required urgent care at the health facility, pregnancy, or any other limitations judged to interfere with study participation or their ability to follow study procedures (cognitive impairment, depression or psychotic disorders). In addition, first degree relatives of participants were excluded.

### Study procedure

The lead author (ERG) and a research nurse trained in the study process implemented the protocol. The research nurse administered the interviewer-assisted questionnaire by reviewing the medical records and conducting the direct interviews, while ERG drew 5mL of venous blood sample from each consenting participant for additional investigation. Participants also submitted spot urine samples. Each questionnaire was issued a unique identifier code to link the participants’ data to the laboratory investigation while maintaining privacy and confidentiality of medical information. The questionnaire comprised demographic information (age and sex), medical history (family history of hypertension and/or diabetes, current hypertension, current diabetes mellitus, current CKD and HIV) and laboratory results (urine dipstick for proteinuria, blood glucose, total cholesterol, low-density lipoprotein, high-density lipoprotein triglycerides, and serum creatinine). The blood pressure, weight and height of each participant was measured by the research nurse in accordance with standard protocols. All the participants with abnormal findings (proteinuria in the urine dipstick, decreased glomerular filtration as well as dyslipidaemia were followed up for further management at the health facility by ERG.

## Measures

Current kidney function was assessed by estimating the serum creatinine and confirming the presence of protein in urine samples. For the detection of proteinuria, spot urine samples were collected from the participants in specimen jars of 40mL. To avoid interpretation bias, the urine test strips were processed in a Urine Analysis Machine Model SLSSUA and interpreted as Protein Negative, Protein trace (15 mg/dL), Protein 1+ (30 mg/dL), Protein 2+ (100 mg/dL) or Protein 3+ (300 mg/dL).

To determine the serum creatinine, 5 mL of venous blood samples were drawn by ERG (a medical doctor) from each participant and sent on ice daily to the Nelson Mandela National Health Laboratory Services for processing, in accordance with standard protocols. The estimated glomerular filtration rate (eGFR) was estimated by using the Chronic Kidney Disease Epidemiology Collaboration Creatinine (CKD-EPI_creatinine_) and the re-expressed four-variable Modified Diet in Renal Disease (MDRD) equations without any adjustment for black ethnicity. There is currently insufficient information in the literature about AmaXhosa people of South Africa and therefore, it is unclear which method is better at detecting kidney damage in all the diverse populations in the country [3,13].

Participants were classified according to the Kidney Disease Outcomes Quality Initiatives CKD classification: GFR categories (ml/min/1.73m^2^); G1 – Normal or High ≥ 90; G2 – Mildly Decreased (60–90); G3a – Mildly to Moderately Decreased (45–59); G3b – Moderately to Severely Decreased (30–44); G4 – Severely Decreased (15–29); G5 – Kidney Failure (≤15).[14] Chronic Kidney Disease (CKD) was defined as an eGFR < 60ml/min/1.73 by CKD-Epi_creatinine_ or the presence of kidney structural abnormalities for more than three months. [14] Hypertension was defined as a persistent elevation of office blood pressure (BP) 140/90mmHg, in two or three readings several hours or days apart, or a personal history of hypertension and/or treatment with anti-hypertensive drugs [15]. Diabetes mellitus (DM) was defined as a persistent elevation of random blood sugar ≥ 11.1 mmol/l on two or more consecutive clinic visits, or a history of diabetes and/or treatment with hypoglycaemic drugs [16].

### Statistical Analysis

Data were analysed with the IBM SPSS Statistics for Windows, Version 27.0 (IBM Corp., Armonk, New York, USA). Descriptive statistics were used to summarise the baseline characteristics and were presented as counts and frequencies for categorical variables, means (± standard deviations) for normally distributed continuous variables, and medians (interquartile range) for non-normally distributed variables. The chi-squared test was used in the bivariate analysis to identify the association between baseline characteristics and CKD. Significant associations between the baseline characteristics and the main outcome (chronic kidney disease) were assessed by applying a multivariate logistic regression model analysis (both unadjusted and adjusted odds ratios) with a 95% confidence interval (95% CI). A p-value of less than 0.05 was considered statistically significant.

## Results

All 389 participants were black Africans, 69,9% of whom were females. The mean age of the participants was 52.3 (±17.5) years. The mean eGFR CK-EPI_creatinine_ was 87,42 (±26.93) ml/min/1.73m*^2^*. The prevalence of CKD (GFR categories G3–G5) was 17.22% (n=67) as determined by CK-EPI_creatinine_, and 17.73% (n=69) as determined by the MDRD equation. Ten participants reported prior diagnosis of CKD at the time of the study (Table 1).

**Table 1:** Demographic and clinical characteristics of the participants.

| Variables | Mean (± Standard Deviation) |
| --- | --- |
| Age (Years) (n=389) | 52.36 ± 17.5 |
| Sex (n=389) |  |
| Female | 272 (69.9) |
| Male | 117 (30.1) |
| Kidney function assessment (n=389) |  |
| Serum Creatinine(μmol/l) | 81.02 ± 36.8 |
| Serum Creatinine(mg/dl) | 0.92 ± 0.41 |
| Urine Proteins (mg/dl) | 8.42 ± 19.32 |
| Estimated Glomerular Filtration rate (ml/min/1.73m<sup>2</sup>) (n=389) |  |
| eGFR CKD-Epi<sub>creatinine</sub> | 87.42 ± 26.93 |
| eGFR CK MDRD | 89.08 ± 38.05 |
| Chronic Kidney Disease (CKD) (n=389) |  |
| eGFR CKD-Epi<sub>creatinine</sub> | 67 (17.22) |
| CKD (eGFR by MDRD) | 69 (17.73) |

CKD-EPI<sub>creatinine</sub>=Chronic Kidney Disease Epidemiology Collaboration Creatinine, MDRD=Modified Diet in Renal Disease, CKD=Chronic kidney disease

### Associations of CKD with potential risk factors

There is a linear relationship between CKD and increasing age; prevalence ranged from 3% at 30–39 years to a maximum of 32.8% at 60–69 years, followed by a decline from 28.4% at 70–79 years to 16.4% at 80 years and over (p-value = 0.000) (**Fig. 1**). There were no sex differences in CKD prevalence (p = 0.4).

**Fig. 1:**
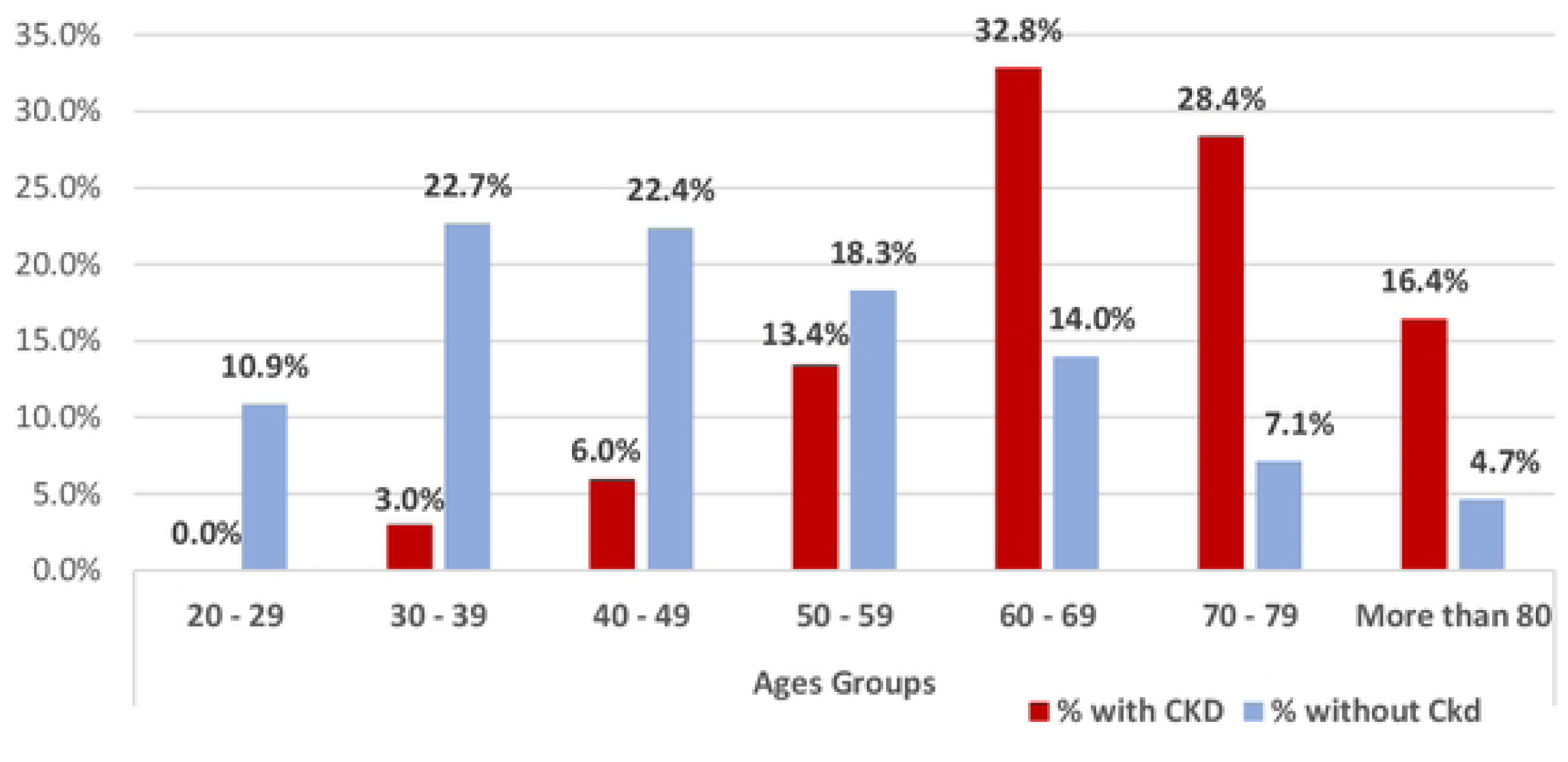
Association between CKD-EPicrealinine and age categories (p=0.000)

In the bivariate analysis, the following potential risk factors were significantly associated with CKD: age (p<0.001), hypertension (p<0.001), diabetes mellitus (p <0.001), HIV (p<0.001) and urine protein (≥ 30mg/dl) (p<0.001) (Table 2).

**Table 2:** Bivariate analysis showing risk factors between participants with and without CKD.

| Variables | CKD | No CKD | Unadjusted Odds Ratios | p-value |
| --- | --- | --- | --- | --- |
| Age | 67.82±12.94 | 49.12±15.99 | 1.08 (1.06 - 1.1) | <.001 |
| Female | 49(73.13) | 223(69.25) | 0.83 (0.46 - 1.49) | 0.529 |
| Hypertension | 60 (89.55) | 144(44.72) | 10.6 (4.7 - 23.89) | <.001 |
| Diabetes mellitus | 24 (35.82) | 56 (17.35) | 2.65 (1.49 - 4.72) | <.001 |
| HIV | 11(16.42) | [18]1(56.21) | 6.54 (3.3 - 12.94) | <.001 |
| Family Hx of hypertension | 16(23.88) | 54(16.77) | 0.64 (0.34 - 1.21) | 0.17 |
| Family Hx of Diabetes mellitus | 10(14.93) | 39(12.11) | 0.79 (0.37 - 1.66) | 0.529 |
| BMI | 24.29 ± 5.27 | 23.76 ± 5.81 | 0.98 (0.94 - 1.03) | 0.489 |
| Urine Protein(≥30mg/dl) | 17.4 ± 30.96 | 6.55 ± 15.29 | 1.02 (1.01 - 1.03) | <.001 |
| Cholesterol | 4.46 ± 0.91 | 4.42 ± 1.12 | 0.96 (0.73 - 1.28) | 0.802 |
| Triglycerides | 1.83 ± 1.01 | 1.59 ± 0.8 | 0.73 (0.44 - 1.23) | 0.236 |

HIV=Human immunodeficiency virus, BMI=Body mass index, CKD=Chronic kidney disease

In the multiple logistic regression analysis, age and presence of proteinuria were significantly associated with the development of CKD in the study cohort. Younger participants were less likely to develop CKD (p<0.001, OR= 0.94, 95% CI 0.91 - 0.96) than older participants.

Similarly, the presence of proteinuria was more likely to be associated with CKD in the study cohort (p 0.002, OR=0.98, 95% CI 0.97 - 0.99) (Table 3).

**Table 3:** Multiple logistic regression analysis of risk factors associated with CKD by CKD- Epicreatinine.

| Variables | Adjusted Odds Ratios (95% CI) | p-value |
| --- | --- | --- |
| Age | 0.94 (0.91 – 0.96) | <.001 |
| Hypertension | 0.48 (0.16 – 1.4) | 0.179 |
| Diabetes mellitus | 0.64 (0.33 – 1.23) | 0.182 |
| HIV | 0.78 (0.3 – 1.98) | 0.595 |
| Urine Protein (≥30mg/dl) | 0.98 (0.97 – 0.99) | 0.002 |

HTN=hypertension; DM=Diabetes mellitus; HIV=Human immunodeficiency virus; eGFR=estimated glomerular filtration rate; CKD-EPI<sub>creatinine</sub>=Chronic Kidney Disease Epidemiology Collaboration Creatinine; OR=Odds ratio.

Thirty-one participants (7.97%) had significant levels of protein in the urine (≥ 30 mg/dl); their mean age was 56.68 (±15.45) years. In the bivariate analysis, the following potential risk factors were significantly associated with proteinuria: hypertension (p = 0.001), HIV (p = 0.045) and elevated Serum Creatinine (µmol/l) (p <0.001). (Table 4).

**Table 4:** Bivariate analysis showing association between potential risk factors and proteinuria.

| Variables | Urine Proteins<br>≥30 mg/dl<br>(n=31) | Unadjusted Odds Ratios | p-value |
| --- | --- | --- | --- |
| Age | 56.68 ± 15.45 | 1.02 (0.99 – 1.04) | 0.141 |
| Female | 22 (5.66) | 1.06 (0.47 – 2.37) | 0.894 |
| Hypertension | 25 (6.43) | 4.17 (1.67 – 10.4) | 0.001 |
| Diabetes mellitus | 9 (2.31) | 1.65 (0.73 – 3.75) | 0.243 |
| HIV | 10 (2.57) | 0.46 (0.21 – 1.01) | 0.045 |
| BMI | 24.21 ± 6.38 | 1.01 (0.95 – 1.08) | 0.707 |
| Serum Creatinine (μmol/l) | 109.16 ± 90.34 | 1.13 (1.09 – 1.17) | <.001 |
| Cholesterol | 4.74 ± 1 | 1.32 (0.91 – 1.92) | 0.148 |
| Triglycerides | 1.56 ± 0.55 | 1.[18] (0.55 - 2.53) | 0.674 |

HIV=Human immunodeficiency virus; BMI=Body mass index. Source: Participants' questionnaire. Data expressed as number (percentage) and mean ± standard deviation. Urine strip results from urine analysis machine model SLSSUA.

In the multiple logistic regression analysis, being hypertensive and having elevated serum creatinine remain significantly associated with the development of proteinuria in the study cohort. Individuals with HTN were four times more likely to develop proteinuria (p<0.015, OR=4.46, 95% CI 1.33 – 14.92) than individuals without HTN, and individuals with elevated serum creatinine were more likely to develop proteinuria (p 0.004, OR=1.01, 95% CI 1 – 1.02) than those without elevated serum creatinine (Table 5).

**Table 5:** Multiple logistic regression analysis of risk factors associated with CKD by proteinuria.

| Variables | Adjusted Odds Ratios | p-value |
| --- | --- | --- |
| Hypertension | 4.46 (1.33 – 14.92) | 0.015 |
| HIV | 1.19 (0.43 – 3.27) | 0.737 |
| Serum Creatinine ($\mu\text{mol/l}$) | 1.01 (1 – 1.02) | 0.004 |

HIV=Human immunodeficiency virus

## Discussion

Despite the increasing prevalence of hypertension, diabetes mellitus and other non-communicable diseases, all known risk factors for the development of CKD in South Africa, the true burden of CKD in the country is unknown. The study reports on the prevalence of CKD and examines some of its known risk factors among the residents of a rural community accessing Mbekweni Community Health Centre in the King Sabata Dalindyebo sub-district municipality of the Eastern Cape province. Findings from this study provide much-needed CKD data on the rural population of South Africa that may inform an effective kidney health prevention programme in the country.

The study found a prevalence of CKD of 17.2% in the sample, with no significant differences shown between the CKD-Epi_creatinine_and MDRD results. This prevalence (17.2%) is higher than the 6.1% reported by Adeniyi et al among a cohort of teachers in an urban area of Cape Town in the Western Cape province in 2017 [9]. It should also be noted that in Adeniyi et al’s (2017) study, there was a considerable difference between the CKD results shown by the two equations; 6.1% according to MDRD and 1.8% according to CKD-EPI_creatinine_ [9]. Similarly, the current study found a much higher prevalence of CKD than the 3.4% found by Peer et al in Cape Town in 2020 [17]. However, the prevalence of CKD in the current study is similar to the 17.3% reported by Matsha et al in Cape Town in 2013 [18]. The wide disparity in the prevalence of CKD can be explained by the peculiarities of each study population. While the participants in the current study were older (mean age 52.36 years) and had a high prevalence of hypertension (52.4%), diabetes mellitus (20.6%) and other cardiovascular risk factors, similar to the participants in Matsha et al in Cape Town [18], relatively younger (mean age 44.1–46.3 years) and more active cohorts, with a lower prevalence of cardiovascular disease, were included in Booysen et al [13], Adeniyi et al [9] and Peer et al [17].

Several studies have compared different glomerular filtration rate equations measured by nuclear medicine methods [13,17,20]. The authors suggest that more studies are needed to validate these equations in the diverse local populations of South Africa. Booysen et al [13] acknowledged that the CKD-Epi_creatinine_ equation may not be as accurate for eGFR assessments among black Africans as it is among the white population. However, this equation represents the most appropriate equation for detecting pre-clinical cardiac and vascular end-organ damage beyond the conventional risk factors found in black Africans [13]. Moreover, Holnes et al [21] reported that both the MDRD and CKD-EPI equations have shown satisfactory accuracy in the South African mixed-ancestry adult population.

In the current study, female participants were predominant (69.9%), which is not surprising, given that this cohort was drawn from the local community health centre. This is in keeping with previous studies in South Africa, where the percentages of female participants were 70.3% [3], 64.1% [17] and 75.2% [18]. However, a study by Kaze et al found a slightly higher male predominance at 53.4% [19]. Although Matsha et al [18] reported a significant relationship between female sex and CKD, the current study found no significant association with sex. This is in agreement with Peer et al [17] and Kaze et al [19], whose studies demonstrated no significant association between sex and CKD. The impact of sex on CKD remains controversial; although some studies document a higher risk of developing CKD among women [18,22,23], others report higher odds among men [24–27].

Previous studies have reported many risk factors for the development of CKD in different population groups in South Africa, especially in the urban parts of Cape Town [9,17,18]. However, such risk factors have not been sufficiently investigated in the rural communities of South Africa, especially in the Eastern Cape province. The current study showed a linear association between CKD and aging population. The mean age of the participants with CKD was 67.82 years, with those above 60 years most affected. This finding corroborates the findings of Peer et al [17] and Kaze et al [19]. However, Adeniyi et al [9] reported the presence of CKD in younger age groups (mean age 47.3 years) among teachers in Cape Town.

The current study found a proteinuria prevalence of 7.97%, which was significantly associated with the presence of hypertension. A similar prevalence of proteinuria (7.2%) and association with hypertension was reported by Kaze et al [19]. However, a lower prevalence of proteinuria (4.5%) in a younger cohort of teachers was reported by Adeniyi et al [9]. It should be pointed out that the urine–protein creatinine ratio was not assessed in the current study, as it was in Adeniyi et al’s study [9]. Although other studies report higher rates of proteinuria among women than men [3,23,28], the current study found no association with sex. In other studies, proteinuria was not evaluated or was related to groups of participants with specific co-morbidities such as HIV ART naïve, HIV on ART, hypertension, or only diabetes mellitus [24, 29–33]

### Study limitations

The study population of the rural KSD sub-district is homogenously black South Africans, which limits the generalisability of the findings to the diverse ethnicity of rural South Africa. In addition, the predominance of females in the cohort reflects a common trend in health service utilisation in South Africa, as elucidated in many population studies in the country [3,17, 18]. Notwithstanding, there was no difference by sex in the CKD prevalence in this study. Future studies on CKD prevalence should target the broader rural population of the country. Furthermore, creatinine was measured using an assay based on the Jaffe reaction, which is more susceptible to interferences than the enzymatic method.

Finally, the use of the CKD-EPI_creatinine_ equation for the detection of CKD needs further validation in the local context, even though both the MDRD and CKD-EPI equations have shown a satisfactory performance in the South African mixed-ancestry adult population. The cross-sectional nature of the study design did not allow for chronicity and might have overestimated chronic kidney disease prevalence in the setting. A previous study showed an overestimation of CKD in an older population and an underestimation in a younger population [34]. Although individuals with probable acute kidney injuries (vomiting, diarrhoea and burns) were excluded from the study, a repeat measurement of creatinine and proteinuria at three months would have established the diagnosis of chronic kidney disease.

## Conclusions

This study found a high prevalence of significant kidney damage (17.2%) and proteinuria (7.2%) in this rural community, largely attributed to advanced age and hypertension, respectively. This finding supports the ideal hospital framework of the national Department of Health in South Africa, which focuses on screening for cardiovascular and other non-communicable diseases at every facility. Early detection of proteinuria and decreased renal function could lead to prompt initiation of preventative measures and management to delay the progression to end-stage kidney failure and mortality.

## Data Availability

The dataset analysed in this study are available with the corresponding author after careful consideration of any written request.

## Acknowledgements

The authors thank the doctors and nurses of the Mbekweni Community Health Centre for their support during the study.

## Authors’ contributions

ERG, GAPE and PY conceptualised, designed and implemented the study protocol. ERG analysed the data and provided the initial draft. OVA and JC critically reviewed and revised the manuscript. All the authors approved the final draft for submission for publication.

